# Harnessing Internet Search Data as a Potential Tool for Medical Diagnosis

**DOI:** 10.1101/2024.05.03.24305695

**Authors:** Gregory J. Downing, Lucas M. Tramontozzi, Jackson Garcia, Emma Villanueva

## Abstract

**Objectives:** To explore the potential and challenges of utilizing internet search data in medical diagnosis, focusing on ethical, technical, and policy considerations by assessing the current state of research, identifying gaps and limitations, and proposing future research directions to advance this emerging field.

**Methods:** A comprehensive analysis of peer-reviewed literature and informational interviews with subject matter experts was conducted to examine the landscape of internet search data utilization in medical research. Searchers were performed for published peer-reviewed literature in PubMed (October to December 2023).

**Results:** Systematic selection according to predefined criteria resulted in the inclusion of 43 articles of the 2,499 identified citations. The analysis revealed a nascent domain of internet search data research in medical diagnosis, characterized by advancements in analytics and data integration. Although significant challenges such as bias, data privacy, and infrastructure limitations hinder widespread adoption, emerging initiatives could reshape data collection methodologies and privacy safeguards.

**Conclusions:** Signals correlating with diagnostic considerations have been identified in certain diseases and conditions, indicating the potential for such data to enhance clinical diagnostic capabilities. However, leveraging internet search data for improved early diagnosis and healthcare outcomes requires effectively addressing ethical, technical, and policy challenges. By fostering interdisciplinary collaboration, advancing infrastructure development, and prioritizing patient engagement and consent, researchers can unlock the transformative potential of internet search data in medical diagnosis to ultimately enhance patient care and advance healthcare practice and policy.

## Introduction

The transition to an era in which information technology (IT) plays a pivotal role in healthcare goes beyond an information engineering advancement to address a substantial medical necessity. Evidence is emerging that internet searches for medical information may help facilitate diagnoses of medical conditions. Machine learning models may predict the diagnosis of a condition more accurately than traditional diagnostic methods. Furthermore, integration of internet search data with a patient’s medical records may provide an opportunity for enhanced screening to identify disease in its early stages. In response to nascent research in this area, the Gordon and Betty Moore Foundation is supporting an initiative to explore the potential to harness internet search data for making medical diagnoses. This report reflects a component of a comprehensive research endeavor focused on addressing pre-hospital diagnostic delays, which encompass the time lapses preceding a patient’s arrival at a healthcare facility where their condition is conclusively diagnosed.^1^

Through a review of the relevant peer-reviewed literature, this report identifies key themes and insights to lay the groundwork for understanding the implications of leveraging internet search data that links with health research datasets resulting in innovative methodologies that empower healthcare professionals to make precise and timely diagnoses.

This work does not consider a patient’s search engine preference for finding health information, nor review the patterns, trends, or accuracy of patient self-diagnosis through internet searches. It does identify the current body of literature from researchers who leverage internet search data to link to other health research data about the individual patient in an attempt to identify a diagnosis. Our objective is to explore the broader landscape of leveraging internet search data in healthcare and its potential for assisting clinicians with diagnoses, and we elucidate promising avenues for researchers to contribute to the enhancement of diagnostic capabilities through thoughtful application of internet search data. In doing so, we sought a nuanced understanding of the possibilities within the realm of healthcare diagnostics with a focus on leveraging search history data to benefit clinical care teams rather than investigating self-diagnosis pathways.

This paper illuminates the research surrounding the potential use of consumer internet search data for early health concern detection, without delving into the clinical validation of such findings. It focuses on the identification of potential diagnostic signals and patterns revealed through this approach to inform the development of predictive models and proactive healthcare interventions, while acknowledging the challenges involved in discerning such signals amidst the vast array of search queries. By leveraging insights from internet search data, healthcare professionals may enhance their ability to identify early warning signs that may lead to timelier interventions and improved patient outcomes.

## Background

Access to accurate medical diagnosis has been hindered by socioeconomic disparities, limited availability of specialized medical professionals, and lack of patient education, among other factors. Inequities in access to high-quality healthcare services exacerbate these challenges, often leading to disparities in health outcomes. Missed or inaccurate diagnoses can lead to delayed or unnecessary treatments, risking worsening of the condition. The historical reliance on direct patient-doctor interactions for diagnosis has often failed to bridge these gaps. The emergence of the internet and digital data in the latter part of the 20th century began to alter this landscape.

Eysenbach highlighted the early potential of the internet in patient education, setting the stage for an ever-increasing reliance on online health information,^2^ but questions remain regarding information accuracy, access and benefits, and privacy.

Internet search data represent one of the largest sources of health data people seek. As of mid- 2023, Google’s daily search volume was over 8.5 billion queries.^3^ Around 5% of Google Searches are health related,^4^ and about 77% of persons with a new diagnosis use search engines.^5^ A recent study showed that 15% of internet searches by individuals with a recent diagnosis involved symptoms of a disease pre-diagnosis,^6^ and 15% of all annual Google Searches are new.^7^

These and other data have prompted a series of research projects to address the feasibility and utility of using internet search data for seeking health services. Although the use of patient search data represents just one facet of technology being explored to help obtain more timely and accurate data about patient conditions,^8^ this paper focuses only on research studies that use internet search data.

### Population Health Research

In population health research, literature is available to assist researchers in approaching the use of internet search data. These studies focus on population health rather than diagnostic search, but remain valuable because they offer methodologies for leveraging internet search data that can benefit research. These studies also delve into how understanding the dynamics of vaccine hesitancy across social media is crucial in devising strategies to promote vaccine acceptance.

Forecasting vaccine hesitancy has become increasingly vital within public health initiatives, and internet search data and social media platforms are pivotal in comprehending the underlying dynamics of this hesitancy.^10^ Leveraging data from search engine logs and social media platforms through machine learning and data analysis provides fresh perspectives on vaccine intentions and behaviors that aid policymakers and healthcare professionals in crafting strategies to tackle vaccine hesitancy.

“Accurate Measures of Vaccination and Concerns of Vaccine Holdouts from Web Search Logs” showcases the potential of utilizing search engine logs for insightful analysis that addresses the public health concerns of patients.^11^ By developing a vaccine intent classifier, researchers accurately detect user searches for COVID-19 vaccines that strongly correlate with Centers for Disease Control and Prevention’s vaccination rates,^11^ enabling real-time estimation of vaccine intent rates across demographics and regions and revealing granular trends in vaccine-seeking behavior.^11^ Machine learning identifies vaccine holdouts, their inclination toward using untrusted news sources, and specific concerns about vaccine requirements, development, and myths.^11^ Understanding these concerns among demographic groups unveils variations in hesitancy, shedding light on those crucial moments when individuals transition from being vaccine holdouts to considering vaccination.^11^

Similarly, the study on COVID-19 vaccine hesitancy and increased internet search queries for fertility side effects following Emergency Use Authorization (EUA) demonstrates the link between public concerns and vaccine uptake.^12^ The surge in fertility-related queries post-EUA, fueled by unfounded scientific claims propagated on social media, underscores the hesitancy regarding potential side effects that influenced vaccine acceptance rates,^12^ emphasizing the importance of addressing specific concerns highlighted by online searches to alleviate hesitancy and promote informed public decision-making.

Research involving empathic engagement with vaccine-hesitant individuals in private Facebook groups highlights the potential for social media platforms to provide a place for health education and discussions.^13^ These moderated discussions positively influenced vaccination intentions and beliefs, representing a promising strategy for combatting vaccine hesitancy.^14^

Social media policies and interventions play a significant role in mitigating vaccine misinformation. Policies implemented by platforms such as Facebook have reduced the reach of anti-vaccine content,^15^ and the systematic appraisal of current social media strategies and their alignment with evidence-based practices represent necessary first steps.^16^ However, the primary focus of these studies involves public sentiment, intentions, and behavioral patterns and not the diagnosis of specific conditions. Leveraging internet search data and social media platforms provides insights into vaccine hesitancy that can drive evidence-based strategies to address hesitancy, promote informed decision-making, and contribute to the success of vaccination campaigns, potentially curbing the spread of vaccine misinformation during public health emergencies.

## Methodology

In addition to conducting interviews with key subject matter experts, we pursued a literature search in PubMed abstract and citation databases based on predefined keyword and term combinations that was performed October 2, 2023, through October 30, 2023. It included a combination of text-words and Medical Subject Headings (MeSH) commonly associated with Google, Bing, Takeout, internet search, web search, search behavior, diagnosis, disease identification, and diagnostic accuracy. Appendix 1 includes a complete list of search terms.

Stringent inclusion criteria were applied to identify relevant studies for analysis according to the PRISMA guidelines. Inclusion was limited to studies that utilized internet search data from Google and Microsoft Bing, which account for more than 90% of all internet searches.^3^ The selected studies’ primary focus was on individual diagnosis and health behavior to ensure a targeted exploration of search data applications in the context of personal health. Studies were required to integrate internet search data with other health research datasets to provide perspective on individual health outcomes and capture the synergistic potential of combining search data with other health-related information.

Exclusion criteria were established to maintain specificity and relevance to the research focus. Studies within the domain of broad population health research were excluded, as was research solely reliant on social media data. These inclusion and exclusion criteria were applied to pinpoint studies aligning with the project’s primary focus: leveraging patients’ internet search data for individual diagnosis and providing patients with information to aid in screening.

All articles retrieved from the initial PubMed search were uploaded to Covidence, where duplicates were removed, and the systematic review process was conducted according to predefined inclusion and exclusion criteria. To reduce errors and bias, the authors independently screened the papers’ titles and abstracts, and full texts of potentially eligible articles were examined for final inclusion. Throughout this process, the authors periodically compared findings, resolving any discrepancies through discussion and consensus to ensure thoroughness and accuracy in study selection.

Table 1 presents the inclusion criteria used to screen publications based on title and abstracts.

**Table 1:** Inclusion Criteria.

| Criteria | Inclusion |
| --- | --- |
| Article type | Peer-reviewed journals. Opinions and commentaries were excluded. |
| Article focus | Use of internet search data, both anonymized and fully identified. |
| Outcomes | The combined use of health research datasets and internet search histories to identify, predict, or confirm a clinical diagnosis. |
| Time | January 1, 2005, to October 30, 2023. |
| Language | English |
| Geography | International |

The exclusion criteria for this systematic review were clearly defined to ensure the relevance and quality of included studies:

- Studies, reports, and publications dated prior to January 1, 2005.
- Articles unavailable in full-text format.
- Articles not written in the English language.
- Newspaper articles, opinions, and commentaries.
- Duplicate studies.
- Studies that did not verify a patient’s clinical diagnosis following analysis of internet search behavior.
- Studies that focused solely on diagnoses at the population level, without specific individual-level data.
- Articles primarily discussing moral, ethical, or privacy considerations related to the use of internet search data without providing analytical insights from the integration of search and clinical data.

## Results

The search initially yielded 6,427 articles, reports, and publications from PubMed. Figure 1 presents the PRISMA flowchart of the record selection process. Duplicates were removed from all articles identified across all searches totaling 3,928 (61% of all results).

**Figure 1.**
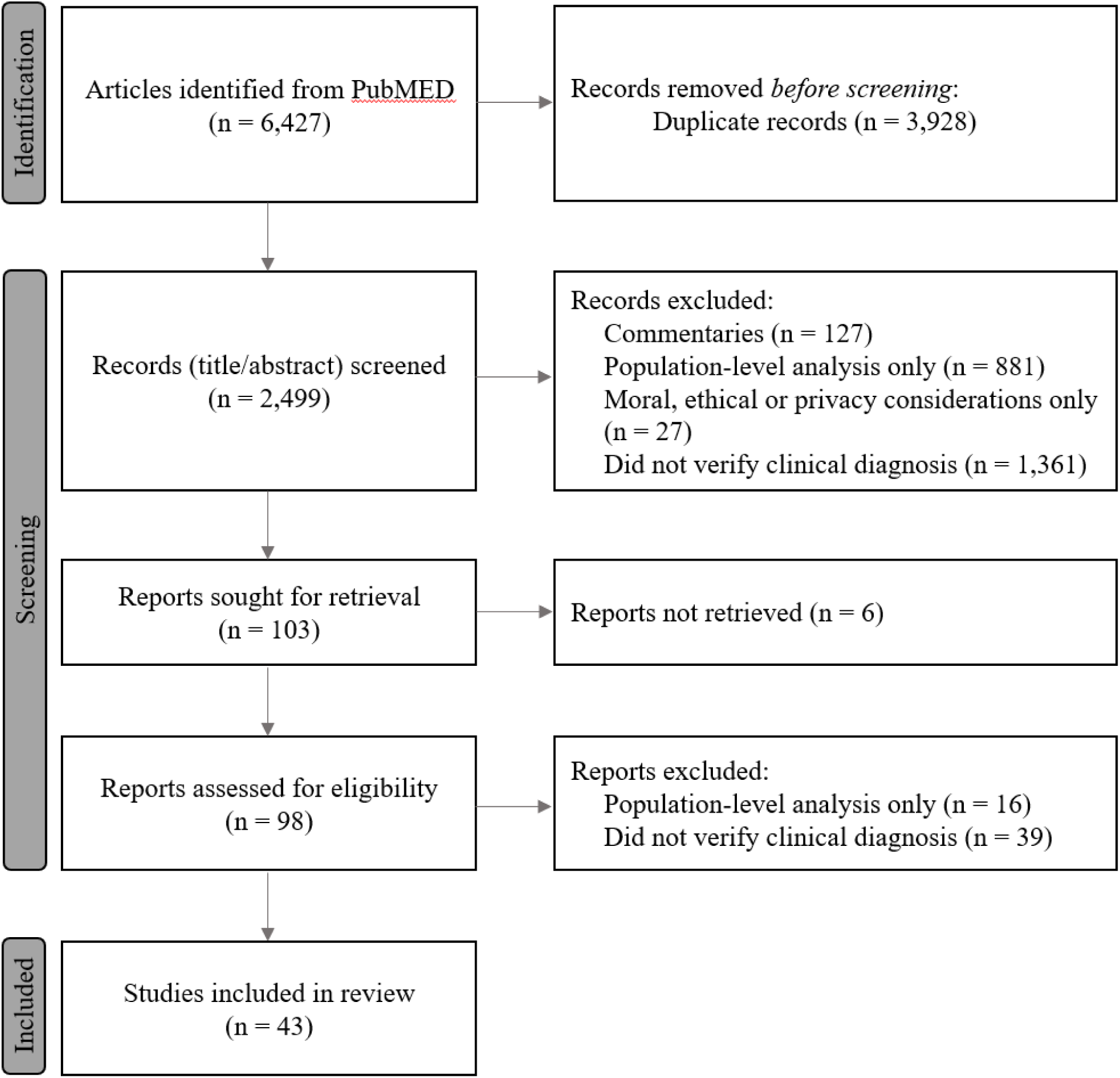
PRISMA Flow for Literature Selection

A total of 2,499 peer-reviewed articles were selected for screening by title and abstract for inclusion and exclusion consideration following the focused criteria. A total of 2,396 articles were excluded based on the following criteria: commentaries (n=127), focused only on population-level disease identification (n=881), focused predominantly on the moral, ethical, or privacy considerations for the use of internet search history while not presenting insights from the analysis of search and clinical data (n=27), or only investigated internet search data without confirming a diagnosis from an independent dataset or from the patient directly (n=1,201).

Full text reports were sought for the remaining 103 articles; however, 6 could not be retrieved. Of the 98 reports obtained, the authors read the full text and excluded 55 reports which focused primarily on population-level analysis (n=16) or which did not confirm a diagnosis from an independent dataset or from the patient directly (n=39). This process resulted in the inclusion of 43 articles in this scoping analysis (see Appendix 2).

### Key Findings

This paper’s primary focus involves exploring research applications of aggregated internet search data, and it offers a nuanced perspective into patient behavior that may unveil potential diagnostic signals that are indicative of various health conditions. The papers identified in the literature review illuminate how internet searches may help in identifying diagnostic signals across a range of diseases and conditions.

Within the realm of health and health services queries, three distinct categories demonstrate generalizability to population-based analyses and individual-specific applications based on search content and patterns. While the use of data from **aggregate, anonymized queries** is widespread, particularly in epidemiological and trending studies at the population level, such use lacks intrinsic value in diagnosing specific conditions for individual patients. These anonymized datasets that lack individuals’ specific informed consent fall under exempt research use. Consequently, this paper does not include detailed examinations of these studies.^18,19^

The second and third categories of research applications using search queries are the focus of this examination. Both include individually consented patient data that may or may not be associated or linked with additional clinical datasets. In the United States, these studies fall under the Common Rule and HIPAA privacy rule.^20^

The second category includes the use of a **search history that can be applied in predicting future search queries** that may strongly correlate with health conditions or disease outcomes. One application of this approach involves developing specialized queries associated with a condition, then searching patients’ internet search logs with that condition and evaluating associated symptoms.^21^ Researchers can then build statistical classifiers that predict future appearances of the landmark queries based on patterns of signals seen in search logs.^21^ Such signals show the possibilities of predicting a forthcoming diagnosis from combinations of subtle temporal signals revealed in searchers’ queries.^21^ This approach was used to establish patient searches of symptoms associated with pancreatic cancer before their clinical diagnosis.^21^

The third category involves the use of peoples’ **internet search logs for which they granted consent for researchers** to access and for which they may have permitted linkage to their health data. Most of these forms of applied research have used retrospective analysis to correlate with features of clinical symptoms or diagnostic tests. In several cases, particularly studies involving behavioral and mental health, prospective associations with Google and Bing search data series have been aligned with clinical outcomes.^22^ Below, we summarize the findings from peer-reviewed publications representing the research conducted on disease or condition diagnosis using the search history to predict future health searches and patient consented data to link to other health research data. Although these data represent a promising avenue for health research, cohort sizes in studies of internet search data linked with clinical records are typically much smaller than those that evaluate only individual internet search data, necessitating careful consideration when interpreting results and designing future studies. Nevertheless, studies that go beyond the use of aggregate, anonymized data offer important insights, particularly in understanding behavioral and mental health conditions.

Each paper’s insights and discoveries are grouped by the health conditions or diseases investigated, allowing for a clear presentation of outcomes and potential diagnostic signals identified across various medical contexts.

### B12 Deficiency

The following case study involves applications of irritable bowel syndrome in the context of public health information, misunderstanding, and patterns of decision-making by individuals. “Evidence From Web-Based Dietary Search Patterns to the Role of B12 Deficiency in Non-Specific Chronic Pain: A Large-Scale Observational Study” used a large dataset of internet search patterns to investigate the relationship between vitamin B12 deficiency and chronic pain.^23^

The study explored the role of vitamin B12 in neuropathy and other neuropsychiatric symptoms using internet search patterns as a proxy for dietary habits.^31^ Researchers analyzed search data from 8.5 million people in the United States, focusing on searches related to food and B12 deficiency symptoms.^23^ Researchers used Bing search data from October 2016 to examine searches for recipes and terms related to chronic pain and B12 deficiency,^23^ then used a linear classification model to link food consumption data with searches for medical terms, finding a strong correlation between food-related search patterns and actual food consumption.^23^ Terms related to neurological disorders were more commonly searched for in conjunction with B12- poor foods.^23^ Also, people who searched for B12-rich foods were less likely to search for medical terms associated with B12 deficiency,^23^ and the average estimated daily B12 consumption for people who inquired about B12 was 2.407 mcg, compared to 2.395 mcg for those who did not, indicating a slight but statistically significant difference.^23^

The study suggests that low vitamin B12 intake may be linked to a broader spectrum of neurological disorders than previously thought.^23^ It emphasizes the potential of using internet search patterns for large-scale health studies.^23^ The researchers recommend further research to explore the clinical significance of these findings and confirm the role of B12 in neuropsychiatric symptoms.^23^ They also note the importance of considering different meat sources in assessing dietary B12 intake.^23^ This study offers insights into the potential use of internet search data in public health research to understand the relationship between diet and disease symptoms.^23^

### Predictive Algorithms for Stroke

Shaklai et al. evaluated the predictive potential of Bing search queries for impending stroke events in an at-risk population in a healthcare setting in Israel.^24^ The study analyzed data from 285 individuals who self-reported a stroke and 1,195 controls, focusing on changes in cognitive traits evident in their internet searches,^24^ and found that certain query attributes related to cognitive function were predictive of an impending stroke.^24^ The model showed high accuracy, particularly as the date of the stroke approached, suggesting that monitoring internet search patterns could offer a valuable tool for early stroke detection.^24^

### Emergency Department Visits

Asch et al. explored the potential of Google Search histories in predicting emergency department (ED) visits and their correlation with clinical conditions.^25^ The study included 103 participants who consented to share their Google Search data collected 7 days before the ED visits; their electronic medical record (EMR) data were included.^25^ The analysis of 591,421 unique search queries revealed that 37,469 (6%) were health related.^25^ In the week before an ED visit, 15% of searches were health related, with many directly related to the participants’ chief complaints.^22^ The study highlights the potential of internet search data in anticipating healthcare utilization and understanding patients’ health-related concerns.^25^

### Intimate Partner Violence

Zaman et al. used Google Search data to identify intimate partner violence (IPV);^26^ 56 participants consented to data analyses that revealed distinctive search characteristics between those with and without IPV experiences,^26^ suggesting that specific patterns in search behavior, including linguistic attributes and search times, can be indicative of IPV.^26^ These findings highlight the potential use of search data for early detection of and intervention in domestic violence.^26^

Youngmann and Yom-Tov analyzed queries from Bing search engine data involving over 50,000 U.S. individuals experiencing IPV.^27^ About half initiated their searches for IPV-related information following an IPV event, while approximately 20% actively concealed their IPV interest. ^27^ Individuals experiencing IPV showed interest in the effects of IPV, seeking help, ways to escape from abusive situations, and more. ^27^ This research suggests that while detecting early signs of IPV through search queries may be challenging, even in the later stages of IPV, interventions such as targeted advertisements to assist people in safely leaving violent situations could be highly beneficial.^27^

### Cancer

“Patterns of Information-Seeking for Cancer on the Internet: An Analysis of Real World Data” was one of the first internet query-based studies to present a detailed analysis of cancer-related internet searches.^28^ It analyzed Yahoo search engine data over 3 months, involving 50,117 users and 225,675 queries.^28^ Findings include a correlation between the aggressiveness of the cancer type and the intensity and duration of the search patterns.^28^ The study employed linear regression and Hidden Markov Models to analyze these patterns^28^ and found a stronger focus on treatment information in searches for aggressive cancers, while support groups were more significant in searches for less aggressive cancers.^28^ This research underscores the potential clinical utility and limitations of using internet search data in understanding the information needs of cancer patients and their acquaintances and suggests that while such data offer valuable insights, they may not represent the diversity of cancer patients’ experiences and needs.^28^

Soldaini and Yom-Tov also demonstrated algorithms that can be designed to identify specific traits of interest in anonymous internet users. The algorithms’ applications in the medical domain demonstrate their effectiveness in identifying potential cancer patients based on search patterns and predicting disease distributions within a population and offer valuable insights for early disease screening and epidemiological studies.^28,29^

#### Parents of Pediatric Oncology Patients

“Health-Related Google Searches Performed by Parents of Pediatric Oncology Patients” analyzed the search behaviors of 98 parents of pediatric cancer patients^31^ and found that parents conducted a higher proportion of health-related searches (13%) compared to the general population (5%).^31^ Searches peaked around key medical events such as diagnosis and treatment phases.^31^

Within health-related searches, 31% involved symptoms, disease, and medical information, and 29% involved hospitals and care sites.^31^ Cancer-specific searches comprised 18% of the health-related queries.^29^ The study emphasized the critical role of the internet in the information-seeking process of parents coping with a child’s cancer diagnosis and treatment and highlighted parents’ significant reliance on the internet for healthcare information in pediatric oncology.^31^ This reliance underscores the need for accessible, reliable online medical information and indicates potential focus areas for healthcare providers in patient and family education.

#### Lung Cancer

“Evaluation of the Feasibility of Screening Patients for Early Signs of Lung Carcinoma in Web Search Logs” explored the use of anonymized web search logs for the early detection of lung carcinoma.^32^ White and Horvitz utilized anonymized search logs from Bing.com involving millions of U.S. English-speaking users.^32^ Of these, 5,443 users who later searched for lung carcinoma symptoms were identified as positive cases.^32^ Statistical classifiers were used to predict search appearances based on earlier search patterns.^32^ Findings showed that certain search behaviors could indicate a higher risk of lung cancer, with true-positive rates ranging from 3% to 57% for different false-positive rates.^32^ The study concluded that web search data could aid in early lung cancer detection, highlighting new directions in identifying risk factors and screening opportunities.^32^

“The Role of Web-Based Health Information in Help-Seeking Behavior Prior to a Diagnosis of Lung Cancer: A Mixed-Methods Study” investigated how online health information influences diagnosis for lung cancer patients.^33^ Through surveys and interviews, the study captured the experiences and behaviors of patients and their next-of-kin.^33^ Quantitative methods were used to establish the proportion of lung cancer cases in which pre-diagnosis web searches occurred.^33^ Qualitative methods were used to explore individuals’ perceptions of the impact their web searches had on the pathway to diagnosis and the barriers that might prevent individuals from accessing the web for information prediagnosis.^33^ Mixed methods were required, because a survey was needed to screen for relevant individuals for interview as low levels of web use among lung cancer patients were expected.^33^ Thus, this study included a cross-sectional, retrospective survey and a qualitative interview study with a subsample of the survey participants.^33^ It found that 20.4% of participants engaged in pre-diagnosis web searches, mainly using Google and NHS Direct.^33^ These searches played a role in all 3 intervals leading to diagnosis: symptom appraisal, decision-making for seeking healthcare, and interaction with health professionals.^33^ The study underscores the growing significance of the internet in early disease detection and patient decision-making.^33^

#### Ovarian Cancer

“Using Online Search Activity for Earlier Detection of Gynaecological Malignancy” focuses on leveraging Google Search data to predict gynecological cancers, particularly ovarian cancer.^34^ This study built upon research by Soldaini and Yom-Tov, which relied on self-identification in queries for outcomes,^35^ while this study employed clinically verified outcomes to enhance the findings’ robustness and reliability. The study, conducted from December 2020 to June 2022 at a London University Hospital, involved 235 women who consented to share their Google Search histories.^34^ It aimed to distinguish between search patterns of women with malignant diseases and those with benign tumors and to explore the possibility of earlier diagnosis through these patterns^34^ and found notable differences in search patterns up to a year before clinical diagnosis, with a predictive model showing an area under the curve (AUC) of 0.82 for individuals who frequently searched for health-related topics,^34^ demonstrating the potential of using online search data as a supplementary tool for early cancer detection.^34^ Chen et al. noted that despite the limited datasets in this study, a tendency is apparent toward heightened online search activity before patients with malignant cases visit a general practitioner.^36^

#### Pancreatic Cancer

“Screening for Pancreatic Adenocarcinoma Using Signals From Web Search Logs” explored the use of Bing web search logs to predict pancreatic adenocarcinoma.^21^ The study involved 9.2 million U.S. English-speaking users, focusing on the feasibility of the early detection of pancreatic cancer by analyzing search patterns.^18^ The researchers analyzed Bing anonymized search logs, looking for patterns that might indicate the early stages of pancreatic adenocarcinoma.^21^ They identified users who searched for symptoms or treatment related to pancreatic cancer and then traced their search history backward, looking for early signals of the disease.^21^ This retrospective analysis searched for distinctive search patterns before the actual diagnosis.^21^ The findings were significant, demonstrating the potential of search log analysis to identify early signs of serious illnesses and that certain search behaviors could be indicative of pancreatic adenocarcinoma, achieving true-positive rates of 5% to 15% with extremely low false-positive rates.^21^ This method could complement traditional diagnostic methods and constitutes an innovative approach suggesting a new direction for cancer screening using web search data in health surveillance and early diagnosis.^21^

### Mental and Behavioral Health

#### Addiction

Nitzburg et al. utilized internet search data to identify patients seeking drug treatment services for alcohol use disorder.^37^ Leveraging internet search data, the study explored how medical symptom queries correlate with subsequent searches about Alcoholics Anonymous and Narcotics Anonymous treatment.^37^ Routine visits to primary care physicians often serve as initial points of contact for problem drinkers, providing an opportunity to motivate them toward alcohol-reduction treatment. Brief intervention (BI) protocols, integrated into routine care, aim to reduce patients’ drinking levels.^37^ By analyzing anonymized Bing search data, the study identified common medical symptoms preceding searches for 12-step programs, illuminating potential avenues to enhance BI’s efficacy in motivating individuals toward seeking treatment.^37^ Findings suggest that emphasizing long-term medical consequences and immediately discomforting symptoms could enhance motivation for seeking treatment. .

#### Anxiety and Depression

The following three studies, focusing on depression and anxiety disorders, present innovative approaches to addressing mental health challenges. Zhang et al. explored the potential of utilizing personal online activity histories from platforms such as Google Search and YouTube to detect depressive disorder among U.S. college students.^38^ By collecting longitudinal data and employing machine learning techniques, the study established correlations between shifts in online behaviors and worsening mental health profiles during the COVID-19 pandemic,^38^ highlighting the feasibility of leveraging ubiquitous online data for noninvasive surveillance of mental health conditions and offering an alternative to traditional screening methods, especially in times of societal disruption.^38^

Zaman et al. expanded the investigation to examine the relationship between changes in Google Search and YouTube engagement behaviors and the exacerbation of depression and anxiety levels among college students during the pandemic.^39^ Through longitudinal data collection and correlation analysis, they identified significant associations between deteriorating mental health profiles and shifts in online behavior and provided insights into the potential use of these behavioral changes to predict mental health conditions.^39^ These findings underscore the importance of utilizing pervasive online data for real-time monitoring and early intervention in mental healthcare, and offer a cost-effective, scalable approach to complement existing screening methods.^39^

In a third study, Zaman et al. proposed an alternative method for identifying individuals with anxiety disorders and estimating their anxiety levels using personal online activity histories from YouTube and Google Search.^40^ By collecting multiple rounds of anonymized data and developing explainable features capturing temporal and contextual aspects of online behaviors, they demonstrated results in detecting anxiety disorders and assessing anxiety levels. This study presents a cost-effective and scalable framework that could be deployed in real-world clinical settings, empower care providers and therapists with insights into anxiety disorders, and enhance mental healthcare delivery.^40^ These three studies highlight how leveraging online data for mental health surveillance and intervention offers new avenues for improving mental health outcomes.

Youngmann et al. revealed that individuals exhibit distinct information-seeking behaviors when using search engines depending on their anxiety level, which is particularly evident in searches for medical symptoms with potentially life-threatening implications.^41^ By analyzing mouse tracking data and other user interactions, a model was developed to predict user anxiety levels that achieved significant correlation with the severity of symptoms searched.^41^ The findings underscore the importance of incorporating user anxiety information to accurately measure search engine performance, which is crucial in effectively delivering critical medical information and suicide prevention resources.

#### Eating Disorders

Sadeh-Sharvit et al. addressed how leveraging internet search data can enable interventions in cases of eating disorders, given their personal and public health costs and the barriers to seeking treatment.^42^ By leveraging internet browsing behavior, the study explored whether data from clinically validated online screens can predict the presence of or high risk for an eating disorder.^42^ Results suggest that a machine learning algorithm incorporating variables such as age, search activity related to eating disorders, and internet usage patterns can identify women screening positive for eating disorders with moderate accuracy, potentially enabling early intervention to reduce the prevalence of these disorders. The study acknowledges the need for larger sample sizes and inclusion of diverse populations, along with the ethical and privacy concerns in implementing predictive models for eating disorder detection using internet browsing data.^42^

#### Mood Disorders and Suicidality

This study conducted at Northwell Health system included 43 individuals ages 15-30 with mood disorders who were hospitalized for suicidal thoughts and behaviors and examined their Google Search activity before hospitalization.^43^ The research identified search patterns related to suicide and behavioral health.^43^ A majority (27/43, 63%) conducted suicide-related searches.^43^ Participants searched for information that matched their chosen method of attempting suicide in 21% (9/43) of cases.^38^ Suicide-related queries also included unusual suicide methods and references to suicide in popular culture.^43^ A majority of participants (33/43, 77%) used queries related to help-seeking themes, including how to find behavioral healthcare.^43^ Queries related to mood and anxiety symptoms were found among 44% (19/43) of participants and included references to panic disorder, inability to focus, feelings of loneliness, and despair.^43^ The results provide insights into digital behaviors of youth with mood disorders facing suicidality, highlighting the potential of internet search data in clinical assessment and intervention strategies.^43^

“Perceived Utility and Characterization of Personal Google Search Histories to Detect Data Patterns Proximal to a Suicide Attempt in Individuals Who Previously Attempted Suicide: Pilot Cohort Study” explored the feasibility and acceptability of using personalized online search data to identify the risk of suicide attempts.^44^ It involved 62 participants with a history of suicide attempts^44^ and analyzed changes in online search behavior up to 60 days before an attempt, revealing patterns such as increased searches related to suicide methods and expressions of anger.^44^ The study highlights the potential of internet search data to identify early warning signs of suicide risk, although participants raised concerns about privacy and accuracy.^44^

#### Psychosis

“Google Search Activity in Early Psychosis: A Qualitative Analysis of Internet Search Query Content in First Episode Psychosis” analyzed Google Search queries of individuals before their first hospitalization for psychosis.^22^ This qualitative evaluation involved 20 participants who provided access to their Google archive data and identified common themes during emerging illness.^22^ Findings revealed that 75% of participants searched for mental health-related information.^22^ 75% of participants included delusions in their queries.^22^ The study concluded that individuals with early psychosis used the internet to understand their symptoms before seeking psychiatric care,^22^ highlighting the potential for tailoring online resources to improve pathways to care and shorten durations of untreated psychoses.^22^

Aref-Adib et al. investigated patterns and consequences of online mental health information-seeking behavior among individuals with psychosis and assessed the acceptability of a mobile mental health application.^45^ Individuals with psychosis commonly seek mental health information online, which proves beneficial when shared with clinicians.^45^ However, when not shared, it can impact healthcare decisions.^45^ The research underscores the need for a collaborative, shared decision-making approach to online health information-seeking that includes discussion with clinicians..^45^ Findings suggest that individuals with psychosis lead active digital lives, indicating that introducing a mental health app into services may be positively received.

#### Schizophrenia

“Utilizing Machine Learning on Internet Search Activity to Support the Diagnostic Process and Relapse Detection in Young Individuals With Early Psychosis: Feasibility Study” explored using internet search data to aid in diagnosing relapses in schizophrenia spectrum disorders (SSD).^46^ It involved 42 participants in the Northwell Health System with SSD and 74 healthy volunteers ages 15-35.^46^ The institutional review board (IRB)-approved study analyzed 32,733 time-stamped search queries.^46^ Machine learning algorithms were developed to distinguish between individuals with SSD and healthy volunteers and to predict psychotic relapses.^46^ Results showed potential for using online search activity as objective data in psychiatric diagnostics and relapse prediction, with classifiers achieving an AUC of 0.74 for diagnosis and an AUC of 0.71 for relapse prediction.^46^ Findings include fewer and shorter searches among SSD participants and specific word use patterns related to symptoms.^46^ This approach represents a novel method for integrating digital data into mental health monitoring and diagnostics.^46^

### Neurodegenerative Diseases

Internet search data have also been used in diagnosing neurodegenerative diseases. Austin et al. explored the relationship between internet search behavior and cognitive function in older adults, with a focus on Alzheimer’s disease.^47^ By continuously tracking and analyzing search terms, the authors found that individuals with poorer cognitive function exhibited distinct patterns in their online searches—they employed fewer unique terms and less common vocabulary.^47^ This suggests that changes in language use during online searches could serve as an early indicator of cognitive decline, thereby potentially enabling treatment before symptoms fully manifest.^47^

Youngmann et al. developed a machine learning algorithm to screen for Parkinson’s disease using data from search engine interactions.^48^ By analyzing the textual content of web queries, the classifier identified individuals at high risk for Parkinson’s.^48^ Longitudinal follow-up revealed that those identified as positive showed a higher rate of progression in disease-related features.^48^ This innovative approach enables large-scale screening for Parkinson’s and offers insights into disease progression, potentially facilitating early intervention and management.

Yom-Tov et al. investigated the potential of internet search engine interactions in identifying individuals with amyotrophic lateral sclerosis (ALS).^49^ By analyzing search engine query data, the authors developed a model capable of accurately distinguishing individuals with ALS from controls and disease mimics.^49^ The prospective validation further supported the approach’s efficacy, indicating its potential as a screening tool to reduce ALS-associated diagnostic delays.^49^ These studies highlight the value of harnessing internet search data for early detection of neurodegenerative diseases, and offer promising avenues for improving clinical outcomes.

### Nutritional and Metabolic Diseases

The utilization of internet search data presents a potential avenue for early detection of nutritional and metabolic diseases such as diabetes. Hochberg et al. analyzed Bing search engine queries from U.S. users to identify symptoms related to diabetes.^50^ Through predictive models, including logistic regression and random forest, the study could distinguish between users diagnosed with diabetes and those querying symptoms associated with diabetes.^50^ The models could detect undiagnosed diabetes patients up to 240 days before they mentioned being diagnosed,^50^ highlighting the potential of utilizing search engine data for earlier diagnosis, which is particularly beneficial for conditions such as type 1 diabetes, where early detection is clinically meaningful.^50^ Additionally, the study suggests the possibility of search engines serving as population-wide screening tools, and hints at potential further improvement by incorporating additional user-provided data.

Lebwohl and Yom-Tov investigated the use of internet search term data to identify symptoms prompting an interest in celiac disease and the gluten-free diet.^51^ By analyzing U.S. Bing search queries, the study characterized symptoms and conditions potentially indicating elevated likelihood of subsequent celiac disease diagnosis.^51^ The study identified various symptoms queried before celiac-related searches, including diarrhea, headache, anxiety, depression, and attention-deficit hyperactivity disorder (ADHD), but the predictive ability of these searches was limited.^51^ The study did observe an increase in antecedent searches for symptoms associated with celiac disease, shedding light on its diverse clinical manifestations and the challenges involved in identifying effective case-finding strategies.^51^ These findings underscore the complex nature of a celiac disease diagnosis and the potential for leveraging internet search data to enhance understanding and detection of such nutritional disorders.

### Cross-Cutting Themes, Lines of Evidence, and Research Gaps

The results of our analysis of the peer-reviewed research for anonymized and non-anonymized research using Microsoft Bing or Google Search data reflect a clearly nascent domain of IT and data research in assisting with diagnosis determinations. Nevertheless, the advances in structured data, large language models (LLMs), powerful data search engines, analytic platforms, and expanding research experiences of health service investigators in population health and individual patient research are promising. Today there is no structured way of designing these types of studies to aid in the diagnosis of diseases and conditions, but among the most visionary applications of search data are those reflected in the development of disease-specific predictive models for classifying internet search terminologies that may one day be applied in real time for clinical decision-making.

The published research addresses feasibility and clinical efficacy (in prospective studies). None of the reviewed studies has addressed clinical utility. However, some of the studies discussed the implications of how the analysis tools and predictive models were used, and some described the conceptualization for the data representations in clinical health record systems.^38–40^ Should this type of research eventually demonstrate clinical utility, one could envision the development of patient applications for empowering individuals; however, the use of internet search data for individual patients has policy and research applications similar to those of other health systems research. A research area that could benefit from population-level applications is rare diseases, where crowdsourcing of queries could be mined for commonalities and integrated with population data, disease registries, and EMRs. Utilities for identifying patient candidates for clinical trial eligibility and enrollment also could be explored.

Researchers publishing results from studies using internet search data are from two general health research domains. Data scientists and research engineers from large technology companies with proprietary technology that supports internet searches have provided methodological innovations in linguistics, mathematics, and information science that open doors for clinical investigations.^27–34, 27–36, 43,46^ Academically oriented health services researchers with experience with large dataset analysis for specific health conditions represent the alternative dimension. The research approaches differ in terms of anonymization, integration with EMRs or other data that enable individual patients to be studied, the size of the groups studied, and the approach to the methods and tools applied. Moreover, it seems likely that fostering research that brings important research questions from the clinical and academic settings together in collaboration with the technology engineering domains would catalyze and accelerate promising clinical and public health insights.

Research questions explored using internet search data usually focus on diseases that evolve over time (subacute or chronic) with a wide array of clinical presentations. One challenge that spans the health domains studied using internet search data for diagnosis in the use of consented, retrospective data involves the substantial opportunities for bias in the methods applied in consenting, patient donation, and other areas. However, the associations of causal effects through statistical analysis and mathematical examinations in population studies that use anonymous data sources can frame insights that can be evaluated through pilot studies and prospective randomized clinical trials that can address or help minimize the effects of bias in patient-provided data.

Several studies integrate datasets from other social media platforms, such as Instagram and X (Twitter), while others use Google Takeout data or Microsoft Bing. We found no publications using Google and Microsoft patient data on the same patients or any studies using the same analytic algorithms. Future work could examine the cross-over effects of patient populations using both data sources because the orientation and structure of the datasets differ.

Only one study to date has used a prospective data collection approach that enables patients to contribute data from the beginning of their enrollment (Katherine Anne Comtois, PhD, University of Washington, personal email correspondence, December 26, 2023). It is unclear whether the search patterns differed in patients who donated their data retrospectively versus patients who donated prospectively. The publications we examined provide no details on the mathematical methods used in classifying terms (there appears to be no consensus or best practice for annotating such data). Thus, reproducing study results may be difficult. We found no publications that have made anonymized research datasets created from their study data available to other researchers for examination. The most detailed descriptive methods publications provide are supplemental data that include search patterns, common terms, and other data classification details. Future research may encourage more open data policies, including the provision of metadata and the descriptive characteristics of the study populations, that would allow others to validate and build on the pilot studies that shape hypothetical associations for detecting and predicting diseases and health conditions.

While cancer diagnosis was the initial clinical domain of disease diagnosis captured in the early literature,^34^ the research has broadened to additional areas, including mental and behavioral health.^35–46^ The ability to obtain search data from patients provides researchers with valuable insights into the patterns of thought, the periodicity of searching patterns, and the thematic aspects of research. Perhaps the most significant domain of search in these studies is in queries that address the patient’s intent to harm oneself or others. A series of studies on integrating patient behavior in social media, online activities, and engagement in risk-taking behavior are underway to evaluate their utility in understanding patient management applications. In these domains, the clinical utility is less focused on diagnosis than on monitoring the patient’s status for management and on using search data as an integral tool to intervene or make therapeutic changes in clinical regimens. Government or non-governmental research organizations are sponsoring several of these studies, marking a milestone for non-industry sponsorship of internet search data application.^1^

From the articles we reviewed and the informational interviews we conducted with researchers with subject matter expertise, there appears to be consensus that assistance with infrastructure development would benefit researchers in designing their studies. In this paper, we have summarized the research findings on tools for harnessing massive datasets and enabling their integration with other datasets, including those with EMR data. We also noted a need for broader information about the nature of the available search datasets, best practices for individuals to manage their datasets with researchers, and the conditions under which their data can be shared. Given the concerns regarding data privacy and security for large datasets in the consumer marketplace and the interplay of these data with HIPAA-regulated data in clinical settings, benefit to the researcher and patient advocacy communities could be achieved by establishing best practices and informational resources to guide future research design, oversight, and patient benefits from the use of their data.

### Researcher Tools

Researchers have created tools to effectively analyze and utilize internet search data and facilitate investigations into internet search studies. This was prompted by the need to comprehensively access and harness the potential of such data. Their integration has eased the identification of early signs of issues, ensured user privacy, and streamlined the investigative process. Innovation in these tools (Appendix 3) often allows researchers to be more successful in their searches, as has been the case in other research domains with novel data sources, such as genomic datasets.

The gTAP Web App prioritizes data privacy and security by allowing participants to download their data without sharing personal account credentials, ensuring a higher level of user trust and confidentiality.^44^ This feature encourages participation in studies involving symptom analysis and diagnostics, fostering collaboration between researchers and users while maintaining data integrity.^44^

LIWC, a text analysis software package, exhibits remarkable potential in differentiating linguistic attributes within search logs.^26^ By identifying linguistic patterns indicating emotional, sexual, or physical abuse, LIWC is instrumental in early symptom identification, providing valuable insights for healthcare professionals and researchers.^26^

The Google NLP API is pivotal in ensuring data privacy and anonymization.^52^ By automatically detecting and removing personally identifying information from search history data before they are saved as research data, this API safeguards the confidentiality of individual study participants,^33^ enabling researchers to delve into symptom analysis and diagnostics using real-world data while upholding ethical standards and privacy regulations.^53^

CrowdTangle, a powerful tool from Meta, aids in monitoring, analyzing, and reporting social media activities.^7^ It effectively offers transparency across various social media platforms, positioning it as an invaluable resource for understanding public discourse and sentiment regarding health-related symptoms and conditions.^7^ It is the most effective transparency tool in the history of social media.^16^

Latent Dirichlet Allocation (LDA) and Differential Language Analysis Toolkit (DLATK) are cutting-edge methodologies in text analysis.^54^ LDA produces clusters of words that occur in the same context across Facebook posts, yielding semantically coherent topics.^16^ DLATK determines the relative frequency with which users employ words (unigrams) and two-word phrases (bigrams)^54^ and can also retain variables and phrases.^54^ Both are pivotal in the identification of potential symptoms or health-related discussions.^54^

In a recent diagnostic study evaluating AI capabilities, the use of the AI chatbot GPT-4 (developed by OpenAI) showcased remarkable proficiency in certain diagnostic scenarios.^55^ Comparing the LLM’s performance with a broad survey of human clinicians, the study revealed that the LLM surpassed human clinicians in accurately determining pretest and posttest probabilities following a negative test result across 5 cases (although performance was comparatively less robust after positive test results).^55^ The study suggests that leveraging probabilistic recommendations from such LLMs could enhance human diagnostic capabilities^55^ and that combining AI’s probabilistic, narrative, and heuristic diagnostic approaches, could contribute to improved diagnostic accuracy through collective intelligence.^55^

These tools offer greater accuracy and prioritize user privacy and data security. Integrating them into research and healthcare systems enables early detection and better understanding of symptoms and contribute to well-being outcomes, especially for older individuals, when combined with a comprehensive support system. As technology evolves, these tools are poised to play an increasingly vital role in enhancing healthcare and advancing diagnostic capabilities.

### Challenges

The use of internet search data to facilitate medical diagnosis faces challenges, including bias, data privacy, and misinformation. The ethical use of patient data is crucial. Wachter and Mittelstadt’s article “A Right to Reasonable Interferences: Re-thinking Data Protection Law in the Age of Big Data and AI” delved into the ethical dilemmas surrounding the use of big data in healthcare,^56^ emphasizing the need to balance patient privacy with the benefits of big data analytics^46^ and the importance of consent and transparency in the collection and use of patient data. It also highlighted biases and inequalities that could arise from mismanaged data practices.^56^ Yom-Tov and Cherlow further emphasized the need to carefully consider the ethical implications and suggest solutions that balance the benefits and challenges of online screening services.^57^

In our exploration of the field of information sciences concerning internet search data, a notable challenge emerged—the distinct lack of infrastructure for constructing a robust analytic approach to leverage these data in medical and health services research. Thus, we investigated alternative open data research organizational models and discovered the pioneering work of Professor Julia Lane. In her book *Democratizing Our Data: A Manifesto*, Lane introduces an organizational model that promises to revolutionize data accessibility and usefulness.^58^ Within this context, the Institute for Research on Innovation and Science (IRIS) stands out with its groundbreaking contribution, the UMETRICS dataset.^58^ UMETRICS constitutes a burgeoning research asset, harnessing administrative data—information collected primarily for administrative purposes, such as billing and record-keeping, that is repurposed for research to analyze healthcare utilization, outcomes, and patterns—from 30 prominent universities that collectively contribute over one-third of federal R&D spending in academia.^58^ This dataset signifies a shift in data practices, fundamentally reshaping data collection methodologies, fortifying privacy safeguards, and fostering the generation of new products.^58^ Notably, IRIS pioneered the inception of “big data” social science research infrastructures.^58^ Central to its mission was confronting the challenge of comprehending the impact of research funding on scientific and economic activities—a formidable task given the complexities of measuring science’s impact.^58^ IRIS responded by spearheading the construction of an entire infrastructure for tracing the effects of research funding on individuals and interconnected networks.^58^ IRIS developed a highly adaptable data infrastructure, composed of a decentralized network of federal agencies responsible for collecting, processing, analyzing, and disseminating data on various aspects of the country, that caters directly to the research university community and provides impactful methods to assess the scientific and economic implications of their research pursuits, thus surpassing the federal statistical system.^58^ Critical to IRIS’s approach was establishing a data infrastructure firmly rooted in transparent governance, robust privacy protocols, and effective confidentiality protections.^58^ This dedication to principled practices was buttressed by a sustainable business model reliant on contributions from data providers and sponsored projects.^58^ This comprehensive approach provides a significant foundation and framework for transformative data activities in the realm of social media and promises accessible and purposeful data utilization^58^

Leveraging the work of such organizations may unify researchers’ approaches in governance, transparency, data sharing, and related aspects essential for utilizing internet search data effectively. Integrating these insights into our analysis could illuminate potential pathways to address critical gaps in this field. We need to establish robust infrastructures that equip researchers with the necessary tools and resources to delve into this type of research at scale. Assessing the true utility of internet search data in medical diagnosis requires comprehensive frameworks that facilitate large-scale analysis while ensuring data privacy and integrity.

Moreover, should research demonstrate the valuable application of these findings, such infrastructures will play a pivotal role in translating discoveries into actionable insights for clinical practice and healthcare policy.

### Healthcare Practice and Policy Implications

The integration of internet search data with health research datasets could hold profound implications for healthcare practice and policy, necessitating careful consideration of both the technical and ethical dimensions. The use of internet search data in healthcare research poses unique challenges that go beyond the scope of traditional regulatory frameworks such as HIPAA. While HIPAA governs the use and disclosure of protected health information held by covered entities, it may not fully address the intricacies of internet search data, which often contain a wealth of information about individuals’ health behaviors and concerns and potentially sensitive details not captured by conventional health records.

In the context of policy implications, IRBs play a crucial role in ensuring ethical research practices and safeguarding participants’ welfare. For research involving internet search data, IRBs must navigate the nuanced landscape of privacy, consent, and potential risks. Unlike conventional clinical data, internet search data may not fall under the strict purview of HIPAA, making it essential for IRBs to establish clear guidelines for these data.

### Research Directions

The findings of this literature review underscore the need for concerted efforts in stimulating research to fully explore the potential clinical utility of integrating internet search data with health research datasets. While our review did not identify a clear clinical utility, it did reveal promising dimensions in behavioral health, early rare disease detection, and cancer diagnoses. The limited amount of research in this domain since the seminal work of White and Horvitz^59,60^ in 2014 and the relative scarcity of research suggest potential barriers related to researchers’ familiarity with the data, technical complexities in mining the data, or other yet-to-be identified obstacles.^61^

To address these gaps and challenges, we propose a multifaceted approach in 4 key areas: First, there is an urgent need to assess the value and utility of internet search and activity datasets in conjunction with health research datasets, including clinical records. This evaluation should explore how such integration can enhance the diagnostic process, contribute to early disease detection, provide personalized health insights, inform data-driven decision-making, and improve overall patient experiences.

Second, future research should focus on mental health, autism, ADHD, and chronic or rare diseases. Tailoring projects to address the unique diagnostic and treatment challenges within these domains may involve the creation of customized algorithms and tools that cater to the needs of these patients and their nuanced health conditions.

Third, the introduction of innovative analytics, including advanced machine learning and AI models, should be a priority. These sophisticated techniques can uncover hidden patterns and trends within the integrated datasets, offering a new frontier in diagnostic accuracy. Developing predictive models could revolutionize healthcare delivery by providing more precise insights into patient conditions and optimizing treatment plans. Furthermore, the advancement of infrastructure platforms that could aggregate search data with other types of online data (social media, generative AI) and clinical data would allow for this research to be conducted at scale and for the introduction of the kind of innovative analytics described above.

Fourth, enhancing patient involvement in modernizing the consent process is paramount. Research should focus on developing innovative strategies that streamline and modernize consent, prioritizing transparency, trust, and patient comfort with the use of their data. Involving patients in shaping research practices ensures ethical, patient-centered healthcare research, reduces administrative burdens, and promotes accessibility and efficiency..

This comprehensive effort aims to propel research in this promising field, overcoming current limitations and paving the way for transformative applications of internet search and activity data in healthcare diagnostics. Innovations, such as the development of reusable platforms for consent and data collection, may improve the engagement of researchers and patients in this research.

Implementing standardized platforms that streamline the consent process and facilitate data collection can significantly enhance research efficiency and scalability. These platforms should incorporate user-friendly interfaces, clear consent language, and robust data security measures to ensure compliance with privacy regulations and promote patient trust. Reusable frameworks can expedite the research process, minimize administrative burdens, and foster collaboration across studies, ultimately advancing our understanding of the clinical utility of internet search data in medical diagnosis.

Respecting patient privacy and obtaining informed consent are foundational principles in healthcare research. Because the integration of internet search data involves potentially sensitive information, careful attention must be paid to ethical considerations. Transparent and user-friendly consent models are needed to ensure that patients understand who will have access to their data and how their data will be used. Innovative approaches to patient engagement should prioritize educating individuals about the benefits and risks of contributing their internet search data to research. Additionally, robust security measures and compliance with privacy regulations are imperative to protect patient confidentiality. Policymakers are pivotal in establishing clear guidelines and regulations that balance the potential benefits of research using internet search data and patients’ medical data with the imperative to uphold patient rights and privacy. Striking the right balance between facilitating research advancements and safeguarding patient interests is critical for the responsible and ethical use of internet search data in healthcare practice and policy.

## Conclusion

In today’s modern healthcare delivery system, many patients remain disadvantaged by the lack of access to timely and accurate diagnosis of disease and health conditions and miss the benefits of early detection and treatment, leading to suboptimal outcomes, health disparities, and ultimately, changes in national economic productivity. Meanwhile, remarkable advances in technical engineering, computing power, social science, data analytics, and information science are leading to unimaginable insights for public health and clinical medicine. Recently, the confluence of these forces in the use of LLMs and generative AI has captured the imagination of the public and health professionals alike.

Initial research studies have illuminated approaches encompassing study design, technical innovations, and data management methodologies tailored to explore the potential utility of and opportunities for leveraging an individual’s internet search data alongside clinical health data to improve early diagnosis of medical conditions. Further research methods are needed to harness the utility of these data in dimensions of case-control studies or small cohorts with detailed associations of disease symptoms and outcomes. Additional studies are needed to validate assumptions made from studies that rely only on search history. Further implementation studies are needed in real-world settings to address the clinical utility of these strategies. Some matters of concern involve the population health costs associated with diagnostic assessments, particularly if the conditions being correlated are of low frequency (or have high false-positive rates) and include substantial medical risk. Today, there is no framework for the clinical adoption of internet search queries in the clinical assessment of patients. For example, how should conditions for a clinical work-up associated with chronic disease concerns be distinguished from those of a rare disease in the use of internet search query applications?

People worldwide use internet search engines and browsers extensively to find health-related information for symptom understanding, self-diagnosis, and self-treatment. The volume of health-related internet searches is immense. An individual’s internet search history is a potentially valuable data source that offers insights into their physical and mental diagnostic journey, leading up to their first healthcare encounter that results in a diagnosis. Such data have enabled researchers to track symptom evolution and even predict medical conditions. Additionally, linking internet search and activity data with healthcare utilization information can unveil disparities in healthcare outcomes based on factors such as insurance type, race, and education. Empowering patients to understand the significance of these data and their utility is essential to enhance their involvement in owning their data and health, thereby driving the potential for improved diagnosis. Nevertheless, despite promising research on this subject, significant epidemiological questions, privacy and consent concerns, questions around technical infrastructure, and the need for further validation and correlation with diagnostic outcomes remain pivotal in advancing this research for the betterment of healthcare.

The Gordon and Betty Moore Foundation’s Diagnostic Excellence Initiative is a step toward a future in which healthcare is more accessible and patient-centric and is driven by IT and data. The field continues to evolve, promising a healthier, more informed society.

The interrogation of internet search data is in its infancy. Initial studies have identified the promise of using internet search data for population- and personal-level health benefits, including assisting in the diagnosis of diseases and conditions, and while the clinical utility of enabling a healthcare professional to apply powerful analytic engines to a specific diagnosis has yet to be attained, research into achieving this goal is accelerating rapidly. This analysis points to the need for strategic and tactical measures to be undertaken by health services researchers, technology engineers, policymakers, and regulators to advance this research for the future and to ensure that the social good of such practices is optimized and that harm and misuse of information are avoided.

## Data Availability

All data produced in the present work are contained in the manuscript

## Acknowledgments

This project was directed by AcademyHealth with funding from the Gordon and Betty Moore Foundation’s Diagnostic Excellence Initiative.

## Reviewers

Henry A. Kautz, PhD, Karen J Maschke, PhD, Elad Yom-Tov, PhD, MA, Chris Riley, JD, PhD, Matthew J Thompson, DPhil, MPH, MBChB

## Appendix 1 Search Term Results

Appendix 1: Search Term Results
| Term | PubMed Results |
| --- | --- |
| "internet search" AND diagnosis | 443 |
| "online search" AND diagnosis | 41 |
| "search engine" AND diagnosis | 124 |
| "web search" AND diagnosis | 13 |
| "search behavior" AND diagnosis | 9 |
| (Google OR Bing) AND diagnosis | 1,566 |
| Takeout AND diagnosis | 4 |
| "internet" AND "early diagnosis" | 40 |
| "internet search" AND disease detection | 39 |
| "online search" AND disease detection | 4 |
| "search engine" AND disease detection | 18 |
| "web search" AND disease detection | 20 |
| "search behavior" AND disease detection | 7 |
| (Google OR Bing) AND disease detection | 580 |
| Takeout AND disease detection | 3 |
| internet AND "disease detection" | 111 |
| "internet search" AND disease identification | 19 |
| "online search" AND disease identification | 31 |
| "search engine" AND disease identification | 124 |
| "web search" AND disease identification | 10 |
| "search behavior" AND disease identification | 1 |
| (Google OR Bing) AND disease identification | 1,194 |
| Takeout AND disease identification | 3 |
| "internet search" AND diagnostic accuracy | 34 |
| "online search" AND diagnostic accuracy | 23 |
| "search engine" AND diagnostic accuracy | 140 |
| "web search" AND diagnostic accuracy | 15 |
| "search behavior" AND diagnostic accuracy | 12 |
| (Google OR Bing) AND diagnostic accuracy | 1,799 |
| Takeout AND diagnostic accuracy | 0 |

## Appendix 2 Literature Matrix

Appendix 2: Literature Matrix
| Title | Brief Synopsis |
| --- | --- |
| **A machine learning algorithm successfully screens for Parkinson's in web users**<br>https://doi.org/10.1002/acn3.50945 | The study aimed to create and assess a novel web-based classifier for Parkinson's disease screening using search engine user data. Applying the classifier to a large cohort identified 1.2% of users over 40 years old as screening positive for Parkinson's, with higher rates in at-risk groups. Longitudinal analysis revealed faster disease progression in individuals classified as positive, highlighting the potential of web-based screening but also raising ethical concerns. |
| **A qualitative study of online mental health information seeking behaviour by those with psychosis**<br>https://doi.org/10.1186/s12888-016-0952-0 | This study delved into the patterns and consequences of online mental health information-seeking behavior among individuals with psychosis and assesses the acceptability of a mobile mental health application (app). Individuals with psychosis commonly seek mental health information online, which proves beneficial when shared collaboratively with clinicians. However, when not shared, it can impact healthcare decisions. The research underscores the necessity for a collaborative approach to online health information seeking, with mental health clinicians encouraging patients to discuss online findings as part of shared decision-making. Findings suggest that individuals with psychosis lead active digital lives, indicating potential positive reception for the introduction of a mental health app into services. |
| **Anxiety and information seeking: evidence from large-scale mouse tracking**<br>https://doi.org/10.1145/3178876.3186156 | The study revealed that individuals exhibit distinct information-seeking behaviors on search engines depending on their level of anxiety and that this is particularly evident in searches for medical symptoms with potentially life-threatening implications. By analyzing mouse tracking data and other user interactions, a model is developed to predict user anxiety levels, achieving significant correlation with the severity of symptoms searched. The findings underscore the importance of incorporating user anxiety information to accurately measure search engine performance. This is particularly crucial in delivering critical medical information and suicide prevention resources effectively. |
| **Assessment of the frequency of online searches for symptoms before diagnosis: analysis of archival data** | This study examined the percentage of people who search for symptoms (on Bing) before they are diagnosed with conditions that have clear physical symptoms. The study found a large variability in the percentage of people who query the internet for their symptoms before a formal medical diagnosis is made. |
| https://doi.org/10.2196/15065 |  |
| **Can internet search engine queries be used to diagnose diabetes? Analysis of archival search data**<br>https://doi.org/10.1007/s00592-019-01350-5 | Some undiagnosed diabetes patients can be detected accurately according to their symptom queries using a search engine. Such earlier diagnosis, especially in cases of type 1 diabetes, could be clinically meaningful. Search engines' ability to serve as a population-wide screening tool could be improved using additional data provided by users. |
| **Combining Traditional and Non-Traditional Data Stream for Understanding Mental Health**<br>https://www.proquest.com/openview/53fa9b9af362636fbfe7dc06abf03ac0/1?pq-origsite=gscholar&cbl=18750&diss=y | This dissertation explored daily online behaviors via Google Search and YouTube platforms to create predictive models for various mental health conditions and introduced a cloud-based framework merging verified mental health indicators with daily online activities, enabling model construction for diverse mental health issues. During a 2-month study involving a college population, Google search logs revealed strong signals identifying individuals with low self-esteem. This mental health assessment framework is cost-effective, time-saving, and scalable, potentially applicable in real-world clinical settings. It enables healthcare providers to non-invasively understand patients' anxiety disorders at any given moment. |
| **Detecting impending stroke from cognitive traits evident in internet searches: analysis of archival data**<br>https://www.jmir.org/2021/5/e27084 | The research suggested that employing algorithms utilizing online search queries could potentially identify populations at risk of stroke and predict near stroke events among those at high risk. Once it undergoes clinical validation, this algorithm holds the promise of facilitating swift preventive measures. Notably, it can be applied cost-effectively, consistently, and across a broad spectrum, with the intention of mitigating stroke events. |
| **Detecting individuals with depressive disorder from personal Google Search and YouTube history logs**<br>https://doi.org/10.48550/arXiv.2010.15670 | This study proposed a personalized framework utilizing Google Search and YouTube engagement logs to detect individuals with depressive disorder, offering a more accessible and timely screening method compared to traditional in-person interviews. |
| **Detecting low self-esteem in youths from web search data**<br>https://doi.org/10.1145/3308558.3313557 | This study addressed the void in examining indicators of low self-esteem, a condition intricately linked to a cycle involving depression and anxiety, at an individual level through the analysis of Google search history data. The research focuses on college students, a demographic susceptible to experiencing depression, anxiety, and low self- |
|  | esteem. They are asked to complete a mental health assessment survey and to provide access to their individual search history. Textual analysis of the search logs reveals prominent patterns capable of identifying individuals currently experiencing low self-esteem. Notably, participants with low self-esteem display fewer searches related to family, friends, and financial aspects. Furthermore, observable differences in the distribution of search categories over time distinguish them from individuals exhibiting moderate to high self-esteem. Leveraging these distinctive markers, the study developed a probabilistic classifier capable of detecting low self-esteem conditions based on search history, achieving an average F1 score of 0.86. |
| **Discovering intimate partner violence from web search history**<br>https://doi.org/10.1016/j.smhl.2020.100161 | This paper proposed an adaptable, lightweight, and widely applicable screening method, validated through authentic data obtained from self-assessment surveys. This technique aims to identify potential indicators of intimate partner violence (IPV) by examining individual Google search histories. Preliminary analysis reveals discernible temporal, textual, and contextual differences in search behaviors between individuals who have or have not encountered IPV. Leveraging these distinctive patterns, a model capable of identifying violence within intimate relationships was constructed, achieving an F1 score of 0.80. Although these findings are preliminary, it is anticipated that this research will stimulate the AI community to address this critical public health issue. Additionally, this study illustrates a clear schematic depicting the processes of consent, data download, and linkage. |
| **Enhancing web search in the medical domain via query clarification**<br>https://doi.org/10.1007/s10791-015-9258-y | This study explored the effectiveness of bridging the gap between layperson and expert medical vocabularies through query clarification, aiming to assist users in finding authoritative and relevant medical information online. Through task-based retrieval studies and the implementation of a supervised classifier to select appropriate synonym mappings, the proposed system demonstrated improved user preference and accuracy in answering medical questions, with up to a 7% increase in correct answers. Additionally, the introduction of the classifier further enhanced the system's performance, resulting in a 12% increase in the fraction of correct answers. |
| **Ethical challenges and opportunities associated with the ability to perform medical screening from interactions with search engines: viewpoint** | The study explored the potential of utilizing search engine logs for screening various medical conditions, offering opportunities for earlier diagnosis and equitable healthcare access but raising ethical concerns regarding privacy and autonomy. It discusses different approaches for providing screening information, from displaying |
| https://doi.org/10.2196/21922 | notices to modifying search results and using advertisements, each with its advantages and disadvantages in terms of privacy, autonomy, and effectiveness. The study emphasizes the need for careful consideration of the ethical implications and suggests solutions that balance the benefits of online screening services with the challenges they pose. |
| **Evaluation of the feasibility of screening patients for early signs of lung carcinoma in web search logs**<br>https://doi.org/10.1001/jamaoncol.2016.4911 | A statistical classifier accurately identified web searchers who later input queries that provided evidence of a recent clinical diagnosis of lung carcinoma. The methods can help identify people at highest risk up to a year in advance of the inferred diagnosis time and identify new risk factors (e.g., house, age, air travel patterns) expressed as evidence in people's search activity and geographic location. |
| **Evidence from web-based dietary search patterns to the role of B12 deficiency in non-specific chronic pain: a large-scale observational study**<br>https://doi.org/10.2196/jmir.8667 | Food-related search patterns were highly correlated with known consumption and food-related searches ($\rho=.69$). Awareness of B12 deficiency was associated with a higher consumption of B12-rich foods and with queries for B12 supplements. Searches for terms related to neurological disorders were correlated with searches for B12-poor foods, in contrast with control terms. Popular medicines, those having fewer indications, and those that are predominantly used to treat pain were more strongly correlated with the ability to predict neuropathic pain queries using the B12 contents of food. |
| **From health search to healthcare: explorations of intention and utilization via query logs and user surveys**<br>https://doi.org/10.1136/amiajnl-2012-001473 | This study aimed to understand the relationship between online health-seeking behaviors and in-world healthcare utilization (HU) by analyzing data from online searches and surveys. By examining search logs and survey responses, the study provides insights into how users transition from online health information seeking to utilizing professional healthcare services. Results suggest a strong correlation between search behavior and healthcare utilization, offering the potential for inferring HU from long-term search patterns without tracking physical location, which could enhance models of user interests and preferences. |
| **From web search to healthcare utilization: privacy-sensitive studies from mobile data**<br>https://doi.org/10.1136/amiajnl-2011-000765 | Aiming to understand the relationship between health information-seeking behavior and engagement with healthcare professionals, this study conducted a privacy-conscious analysis of geo-tagged data from mobile devices. By analyzing anonymized logs of mobile interactions focusing on search queries and distances to medical care centers, the study investigated the sequence of health-related searches leading to |
|  | observed healthcare utilization indicators. Results revealed that the duration between symptom searches and evidence of healthcare utilization varies depending on symptom severity. Statistical models were developed to predict forthcoming healthcare utilization that achieved predictive accuracies ranging from 65% to 90%. The study emphasizes the importance of privacy-sensitive analysis in generating insights into health information seeking and healthcare engagement, highlighting the potential of large-scale mobile device studies in understanding individuals' pathways to professional medical care. |
| **Google search activity in early psychosis: a qualitative analysis of internet search query content in first episode psychosis**<br>https://doi.org/10.2196/19348 | The study examined the Google search histories of people experiencing their initial episode of psychosis before their initial hospital admission to uncover recurring topics and subjects they were exploring during the onset of their illness. It seems that individuals in the early stages of psychosis are turning to the internet to seek information about their initial symptoms and encounters before seeking psychiatric assistance. Enhancing our comprehension of how individuals in the early phases of psychosis search online for information about their experiences could assist mental health professionals in customizing online resources. This customization could enhance access to care and potentially decrease the duration between the onset of symptoms and the initiation of appropriate treatment for psychosis. |
| **Google search histories of patients presenting to an emergency department: an observational study**<br>https://doi.org/10.1136/bmjopen-2018-024791 | The aim of the study was to assess patients' readiness to share and connect their previous Google search records with information from their electronic medical records (EMRs, and to investigate correlations between search histories and clinical conditions. Out of all searches made within 7 days before an emergency department (ED) visit, 5% were related to health. Among the participants who utilized Google in the week leading up to their visit to the ED, 53% sought information directly linked to their primary health concern. The fluctuations in both the quantity and substance of search activity before an ED visit indicate that there are possibilities to predict and enhance healthcare utilization before such visits occur. Also, patients showed a willingness to grant researchers simultaneous access to their Google search histories and EMR data. |
| **Googling for a diagnosis—use of Google as a diagnostic aid: internet based study** | Google searches revealed the correct diagnosis in 15 (58%, 95% confidence interval 38% to 77%) cases. As internet access becomes more readily available in outpatient |
| https://doi.org/10.1136/bmj.39003.640567.ae | clinics and hospital wards, the web is rapidly becoming an important clinical tool for doctors. The use of web-based searching may help doctors diagnose difficult cases. |
| **Health-related Google searches performed by parents of pediatric oncology patients**<br>https://doi.org/10.1002/pbc.27795 | The content found in Google searches can provide valuable insight into the concerns of parents with children diagnosed with cancer. Analyzing this content could guide us toward more inclusive strategies for educating and supporting families. The study's goal was to pinpoint the priorities and areas where parents lacked information before and after the diagnosis. Searches related to health surged in the months leading up to the child's cancer diagnosis, predominantly focusing on symptoms and logistical information. Following the cancer diagnosis, health-related searches reached their peak after about a month, and these searches included general health inquiries alongside specific cancer-related searches. |
| **Identification of pathological types of adnexal masses from ultrasound images using deep learning model**<br>https://doi.org/10.1002/uog.25071 | This research assessed deep-learning models to distinguish between benign, malignant, and borderline adnexal masses. Additionally, the study aimed to classify 15 pathological types based on ultrasound images. This ongoing preliminary investigation has indicated the acceptability and feasibility of examining online search behaviors among women with gynecological symptoms. Despite the limited dataset, there seems to be a tendency toward heightened online search activity before patients with malignant cases visit a general practitioner. Future efforts will concentrate on expanding the dataset to facilitate the application of machine learning techniques for understanding trends in online search patterns. The goal is to develop a classification model capable of providing early indications for identifying malignant gynecological diagnoses. |
| **Identifying amyotrophic lateral sclerosis through interactions with an internet search engine**<br>https://doi.org/10.1002/mus.27991 | The study investigated the feasibility of using internet search engine interactions to identify individuals with amyotrophic lateral sclerosis (ALS), with the goal of reducing the time from symptom onset to diagnosis. Through analysis of search engine query data from ALS patients and control groups, the study developed a model that distinguishes between ALS cases and controls with promising accuracy (AUC of 0.81). Prospective validation supports the potential of search engine interactions as a screening tool for ALS, although distinguishing ALS from disease mimics remains challenging. These findings underscore the need for further research to explore the role of search engine data in ALS diagnosis and in reducing diagnostic delays. |
| **Identifying sleep disorders from search engine activity: combining user-generated data with a clinically validated questionnaire**<br>https://doi.org/10.2196/41288 | This study explored the potential of utilizing search engine activity alongside a web-based sleep questionnaire to conduct wide-scale screening for common sleep disorders. By analyzing data from 397 participants who completed the questionnaire, the study demonstrated that diurnal patterns of individuals with sleep disorders are shifted by 2 to 3 hours compared to controls. While search engine data alone may not suffice for screening, targeted advertisements coupled with web-based tools could aid in early detection and encourage individuals to seek further assessment for sleep disorders. |
| **Individual-level anxiety detection and prediction from longitudinal YouTube and Google search engagement logs**<br>https://doi.org/10.48550/arXiv.2007.00613 | The paper introduced an innovative method aimed at identifying individuals experiencing anxiety and assessing the severity of their anxiety levels by analyzing their personal online activity histories obtained from popular platforms such as YouTube and Google Search. These platforms serve millions of users daily. The study conducted a longitudinal analysis, gathering multiple sets of anonymized YouTube and Google Search logs from volunteers, alongside clinically validated anxiety assessment scores. Subsequently, the researchers devised distinct features capturing the temporal and contextual aspects of online behaviors. Utilizing these features, the team trained models capable of (i) detecting individuals with anxiety disorder, achieving an average F1 score of $0.83 \pm 0.09$, and (ii) estimating anxiety levels by predicting the widely accepted Generalized Anxiety Disorder 7-item scores (ranging from 0 to 21) with a mean square error of $1.87 \pm 0.15$, leveraging ubiquitous individual-level online engagement data. The proposed framework for assessing anxiety proves to be cost-effective, time-efficient, and scalable, and it holds promise for real-world implementation in clinical settings. This approach enables healthcare providers and therapists to non-invasively understand patients' anxiety disorders at any given moment, providing valuable insights for personalized care. |
| **Inferring individual attributes from search engine queries and auxiliary information**<br>https://doi.org/10.1145/3038912.3052629 | The study introduced an algorithm designed to identify specific traits of interest in anonymous internet users, which is essential for conducting research on various human behaviors including medical conditions. By leveraging labeled examples and statistical data about the population, the algorithm can accurately assign labels to unseen examples, facilitating research in domains where direct identification is challenging due to privacy concerns. The algorithm's applications in the medical domain demonstrate its effectiveness in identifying potential cancer patients based on |
|  | search patterns and in predicting disease distributions within a population, offering valuable insights for early disease screening and epidemiological studies. |
| **Internet search activity of young people with mood disorders who are hospitalized for suicidal thoughts and behaviors: qualitative study of Google search activity**<br>https://mental.jmir.org/2021/10/e28262 | This study found that a significant proportion (27 out of 43, equivalent to 63%) of the participants engaged in searches related to suicide. A deeper comprehension of the searching behavior among individuals contemplating suicide can enhance strategies for outreach, evaluation, and intervention for those at risk. The utilization of search data could also offer advantages in the continuous care provided to individuals dealing with suicidal thoughts or tendencies. |
| **Internet searches and their relationship to cognitive function in older adults: cross-sectional analysis**<br>https://doi.org/10.2196/jmir.7671 | Internet searches were correlated with cognitive decline. The results suggest that early decline in cognitive function may be detected from the terms people search for when they use the Internet. By continuously tracking the basic aspects of Internet search terms, it may be possible to detect cognitive decline earlier than currently possible, thereby enabling proactive treatment and intervention. |
| **Internet searches for medical symptoms before seeking information on 12-step addiction treatment programs: a web-search log analysis**<br>https://doi.org/10.2196/10946 | This study examined the search patterns of people prior to treatment for addiction. The results suggest that many common or non-severe medical symptoms and conditions motivate subsequent interest in addiction prevention programs. In addition to highlighting severe long-term consequences, brief interventions could be restructured to highlight how increasing substance misuse can worsen discomfort from common medical symptoms in the short term, as well as how these worsening symptoms could exacerbate social embarrassment or decrease physical attractiveness. |
| **Intimate partner violence as reflected in Internet search data**<br>https://doi.org/10.1177/08944393221084074 | Queries from Bing search engine data of more than 50,000 US-based individuals suffering from IPV were extracted and analyzed. Approximately half of the users begin to search for IPV following an acute event (physical violence or abuse), and 20% of users actively hide their interest in IPV. The topics of interest to people who experience IPV include the effects of IPV, help-seeking, and methods to escape from IPV. Early cues of IPV may be difficult to detect within search queries, and in the late stage in which many IPV users are identified, interventions such as ads to guide people to safely exit violent situations could be beneficial. |
| **Patterns of information-seeking for cancer on the Internet: an analysis of real world data**<br>https://doi.org/10.1371/journal.pone.0045921 | This study investigated the searches people made on Yahoo search following their cancer diagnosis or a cancer diagnosis of their acquaintances. It shows that search data can be used to investigate medical questions on a large scale. By understanding the patterns of internet use, physicians can use this tool as a powerful partner rather than a source of distress in the care of their patients. Moreover, internet content providers need to personalize their content by taking patient search history into account, because, as the findings demonstrate, information needs change over time. |
| **Perceived utility and characterization of personal Google search histories to detect data patterns proximal to a suicide attempt in individuals who previously attempted suicide: pilot cohort study**<br>https://doi.org/10.2196/27918 | The aim in conducting this research was to assess the viability and approval of utilizing individualized online information-seeking actions to identify the likelihood of suicide attempts. Variations in online search habits could serve as a viable and permissible method for detecting the risk of suicide. A personalized examination of online information-seeking conduct revealed significant alterations in search behaviors and search phrases associated with early indicators of suicide, noticeable within the period spanning from 2 months to 7 days prior to a suicide attempt. |
| **Predicting eating disorders from Internet activity**<br>https://doi.org/10.1002/eat.23338 | The algorithm built from internet activity reached an accuracy of 52.6% in predicting eating disorder risk/diagnostic status. The most predictive internet search history variables were the following: use of keywords related to eating disorder symptoms and websites promoting eating disorder content, participant age, median browsing events per day, and fraction of daily activity at noon. |
| **Screening for cancer using a learning Internet advertising system**<br>https://doi.org/10.1145/3373720 | The study demonstrated the effectiveness of using online advertising systems such as Bing and Google ads in identifying individuals who may have symptoms consistent with suspected cancer. By providing clinically verified questionnaires and analyzing responses, a classifier trained on past Bing queries achieved a predictive accuracy of 0.64 for suspected cancer. Furthermore, leveraging questionnaire responses within Google's advertisement system enabled the identification of individuals likely to have suspected cancer, highlighting the potential of modern advertising platforms to aid in the early detection of serious medical conditions. |
| **Screening for pancreatic adenocarcinoma using signals from Web search logs: feasibility study and results**<br>https://doi.org/10.1200/JOP.2015.010504 | Signals in search logs show the possibilities of predicting a forthcoming diagnosis of pancreatic adenocarcinoma from combinations of subtle temporal signals revealed in the queries of searchers. |
| **Seeking insights about cycling mood disorders via anonymized search logs**<br>https://doi.org/10.2196/jmir.2664 | This study examined Bing searches of people with cycling mood disorders. |
| **Symptoms prompting interest in celiac disease and the gluten-free diet: analysis of internet search term data**<br>https://doi.org/10.2196/13082 | An examination of Bing searches related to celiac disease found an increase in antecedent searches for symptoms known to be associated with celiac disease, a rise in searches for depression and anxiety, and an increase in symptoms that are associated with celiac disease but may not be reported to healthcare providers. The protean clinical manifestations of celiac disease are reflected in the diffuse nature of antecedent internet queries of those interested in celiac disease, underscoring the challenge of effective case-finding strategies. |
| **The relationships of deteriorating depression and anxiety with longitudinal behavioral changes in Google and YouTube use during COVID-19: observational study**<br>https://doi.org/10.2196/24012 | This study investigated the relationship between changes in Google search and YouTube engagement behaviors and the exacerbation of depression and anxiety levels among college students during the COVID-19 pandemic. Through longitudinal data collection and correlation analysis, the study identified significant associations between deteriorating mental health profiles and shifts in online behavior, suggesting the potential utility of these behavioral changes as predictive indicators of mental health conditions. |
| **The role of web-based health information in help-seeking behavior prior to a diagnosis of lung cancer: a mixed-methods study**<br>https://doi.org/10.2196/jmir.6336 | This study investigated the role of web-based information in the pathway to diagnosis for lung cancer patients. Findings indicated that although only a minority (20.4%) reported using the web before diagnosis, both patients and their next-of-kin perceived its impact across all intervals of the diagnostic pathway. The study suggests that while the current role of the web in pre-diagnosis is limited, it holds potential for reducing |
|  | delays in diagnosis, especially as technology familiarity increases, and proposes the division of the diagnostic interval into two subintervals for future exploration. |
| **Using online search activity for earlier detection of gynaecological malignancy**<br>https://doi.org/10.21203/rs.3.rs-2765605/v1 | This study examined the potential of online search data to detect gynaecological cancer in individuals with confirmed diagnoses, suggesting that differences in search patterns were noticeable as early as 360 days before primary care referral. Using a classification model, the study achieved its highest accuracy in predicting cancer risk using data from 60 days before referral, particularly in individuals who frequently searched for health-related topics online. The findings suggest that online search data could offer personalized risk profiles for gynaecological cancer, offering a complementary approach to conventional screening methods and potentially aiding in the earlier detection of various conditions, including cancer. |
| **Utilizing machine learning on internet search activity to support the diagnostic process and relapse detection in young individuals with early psychosis: feasibility study**<br>https://doi.org/10.2196/19348 | The primary objective of this study was to create computational algorithms utilizing internet search patterns, aiming to aid diagnostic processes and recognize potential relapses among individuals diagnosed with schizophrenia spectrum disorders. The research revealed discernible distinctions in the timing, frequency, and nature of online search behavior among young individuals (ages 15-35) with schizophrenia spectrum disorders when compared to their healthy counterparts. Furthermore, alterations in language use and behavioral patterns were detected in the month preceding a relapse leading to hospitalization in individuals diagnosed with schizophrenia spectrum disorders. The study suggests that online search activity holds potential as a means to gather objective and easily accessible markers of psychiatric symptoms. The integration of search behavior as supplementary information related to behavioral health could represent a significant advancement in utilizing unbiased digital data to enhance the monitoring of mental health. |

## Appendix 3 Tools Developed to Assist Researchers in the Use of Search Data

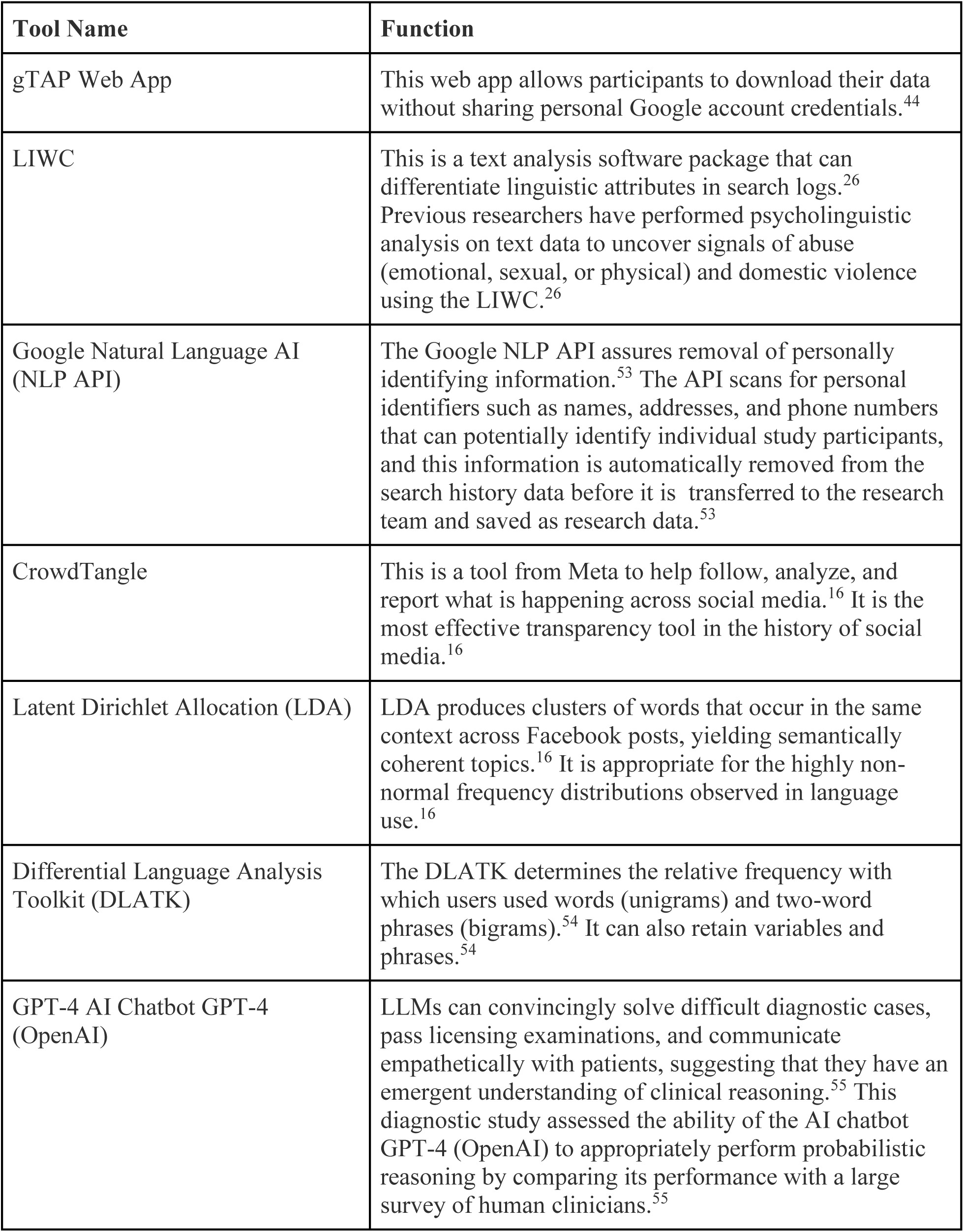

